# Efficacy and safety of SP16 in preventing Acute Kidney Injury in at-risk subjects with chronic kidney disease undergoing elective cardiac surgery using the heart-lung-machine (EASE-AKI): study protocol for a prospective, randomised, double-blind, placebo-controlled clinical trial

**DOI:** 10.64898/2026.06.11.26355378

**Authors:** Tilman Jobst-Schwan, Karl Bihlmaier, Dana Austin, Cohava Gelber, Robert Cesnjevar, Frank Harig, Mario Schiffer

## Abstract

**Background:** Cardiac surgery using cardiopulmonary bypass uses controlled hypoperfusion which leads to relative organ damage. Acute kidney injury is the most frequent and most important organ failure, in particular in patients with chronic kidney disease. To date, there are no approved drug treatments that could effectively prevent acute kidney injury. SP16, an agonist of the low-density lipoprotein receptor-related protein 1, has been shown to exert both reno- and cardioprotective effects in preclinical trials. Early clinical use of SP16 in phase I trials was safe. Administration of SP16 had beneficial trends on inflammatory response and infarct size in patients with ST-segment elevation myocardial infarction. The primary objective of this phase IIa trial is to demonstrate that injection of SP16 is safe and superior to placebo in preventing cardiac surgery-associated acute kidney injury within 7 days after surgery.

**Methods:** This randomised, double-blinded, placebo-controlled, single centre study evaluates the efficacy and safety of SP16 in 120 high-risk chronic kidney disease patients with disease stadium G2-G3b undergoing cardiac surgery who are randomised into one of two treatment groups in a 1:1 ratio: SP16 (12 mg) or placebo. The study medication is administered via two subcutaneous injections, with the first dose given before surgery, followed by an additional dose after 9 h. Primary endpoints are the incidence of acute kidney injury during 7 days post-surgery and the frequency of adverse events within 72 h after index surgery. Important secondary endpoints include the incidence of major adverse kidney events at day 90 and impact on cardiac function. Safety assessments encompass adverse events, vital signs, electrocardiograms and routine safety laboratory tests. Additional evaluations include pharmacokinetics and immunological biomarkers.

**Discussion:** This single-centre phase IIa trial will assess the incidence of cardiac surgery-associated acute kidney injury, describing the renoprotective potential of SP16 and its safety profile in patients undergoing cardiac surgery.

**Trial registration:** EUCT, EUCT number 2025-522491-89-00. Registered February 2, 2026, https://euclinicaltrials.eu/ctis-public/view/2025-522491-89-00, Patient recruitment is planned to start April 1, 2026.

**Structured summary {1b}:** 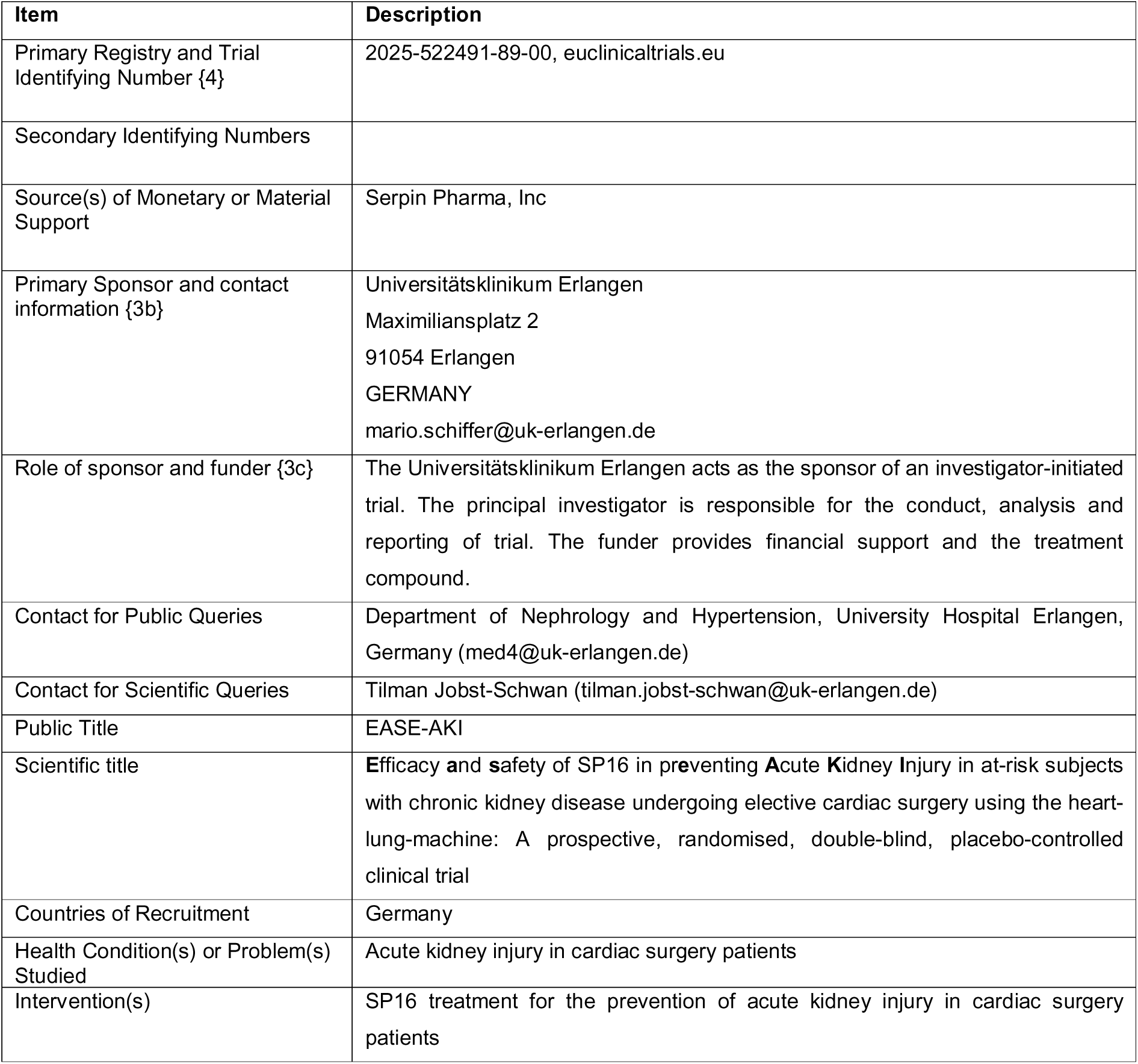

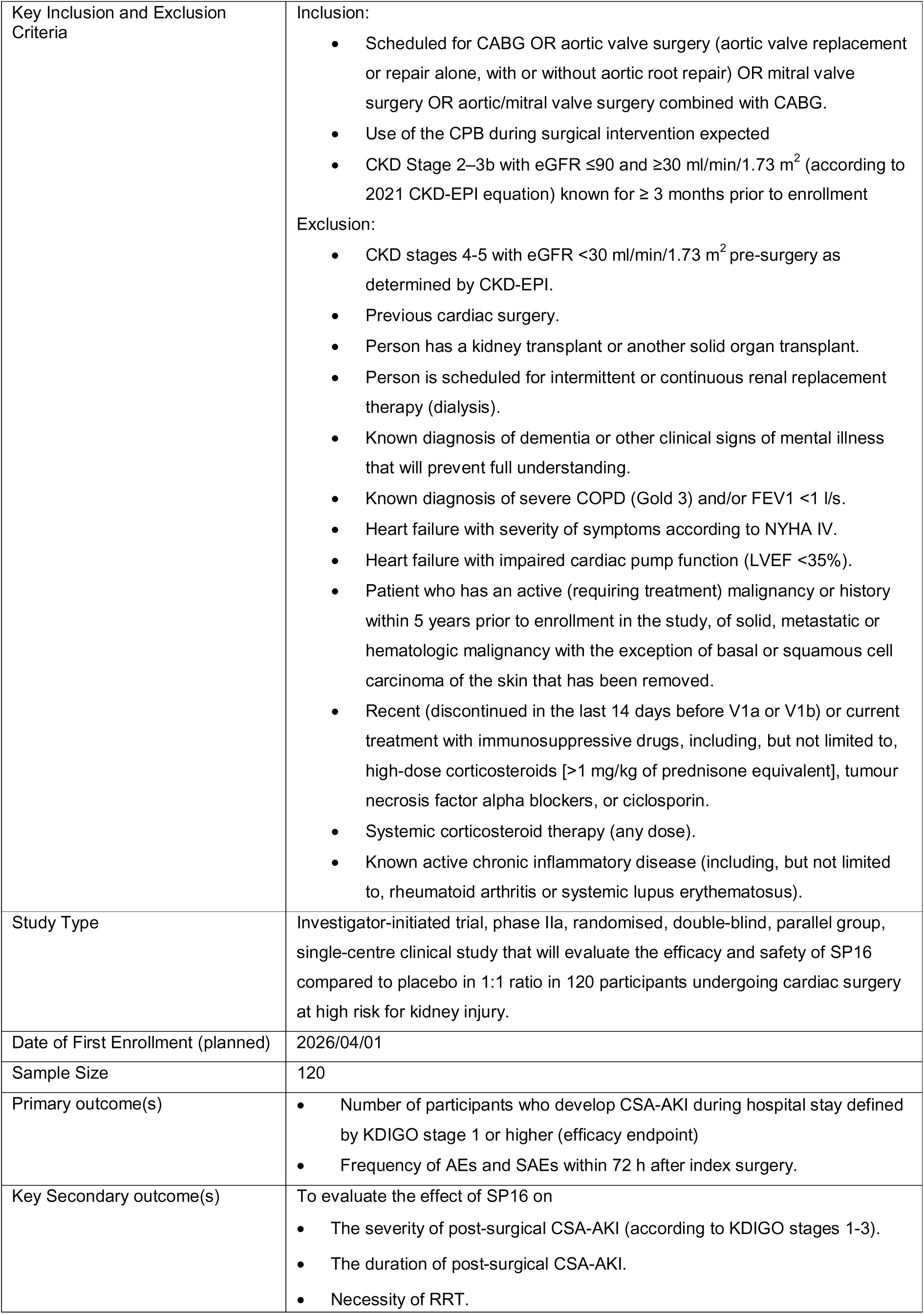

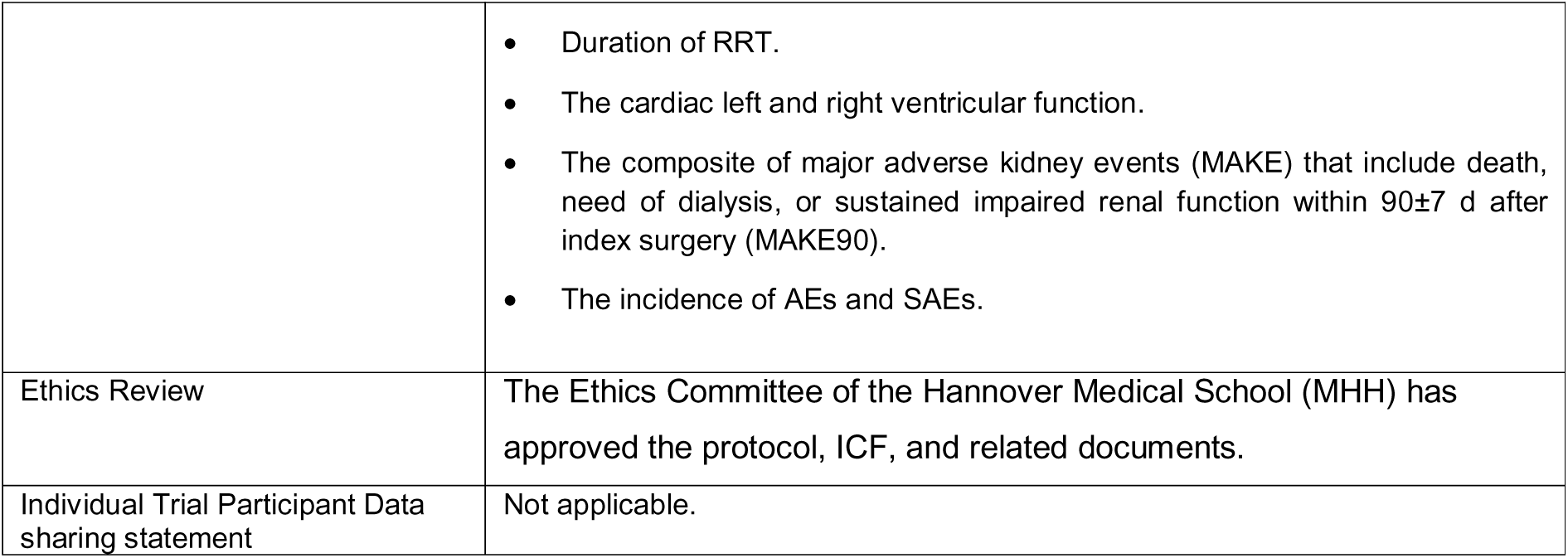

**Protocol version {3}:** 2026 February 4; version 1.1

## Introduction

### Background and rationale {9a}

Cardiac surgery (CS) using cardiopulmonal bypass (CPB) leads to acute kidney injury (AKI) in about 19-45% of patients (1–6). The rate is elevated to 50% in patients with existing chronic kidney disease (CKD) (7). Severe AKI requiring renal replacement therapy (RRT) occurs in 2-6% of these patients (3–6). In addition, CS-associated AKI (CSA-AKI) is significantly associated with the induction of CKD or progression to CKD (Coca et al., 2012), increased rates of heart failure events, and death (8, 9). Even after full recovery of renal function, long-term survival is decreased (10). Although the optimisation of supportive care according to the KDIGO guidelines is beneficial, the rate of CSA-AKI is still high (55.1 vs. 71.7%) (11). KDIGO guidelines (“KDIGO CT surgery bundle”) consist of the following measures: avoidance of nephrotoxic agents, discontinuation of angiotensin converting enzyme inhibitor (ACEi) and angiotensin II receptor-1 blockers (ARBs) for the first 48 h after surgery, close monitoring of serum creatinine and urine output, avoidance of hyperglycemia for the first 72 h after surgery, consideration of alternatives to radiocontrast agents, close hemodynamic monitoring using a PiCCO analysis with an optimisation of volume status and hemodynamic parameters according to a prespecified algorithm: Stroke volume variation (SVV) <11% (otherwise therapy with 500-1000 ml crystalloids), cardiac index (CI) >3 l/min/m^2^ (otherwise therapy with dobutamine or epinephrine), mean arterial pressure (MAP) >65 mmHg (otherwise therapy with norepinephrine). Additionally, even minor increases in serum creatinine post-surgery are linked to a significant rise in mortality among CS patients (12, 13). The occurrence of AKI in intensive care unit (ICU) patients also leads to increased direct and indirect healthcare costs and resource utilisation.

Despite the critical importance of CSA-AKI, there is currently no reliable evidence supporting specific interventions to prevent CSA-AKI. In 2013, the Cochrane Collaboration concluded that none of the existing interventions reported effectively protect the kidneys during the perioperative phase, nor do they appear to cause increased harm. Several pharmacological strategies have been investigated for the prevention of CSA-AKI without significant benefits (14–21). Recently, intravenously administered amino acids in CS patients was shown to be renoprotective, but the AKI rate remained high (26,9% vs. 31,7%) (1). Thus, there is still an urgent need for renal protection in CS patients.

Upon AKI, Wnt/β-catenin signaling is upregulated in kidney tubules as protective mechanism (22). Low-density lipoprotein receptor-related proteins (LRPs) are members of the low-density lipoprotein (LDL) receptor family that are involved in endocytosis but also in complex signaling processes (Fig. 1). The ubiquitously expressed LRP1 protein contributes to Wnt/β-catenin signaling and cholesterol metabolism, for example in macrophages and T-cells. LRP1 upregulates Wnt5a that reduces intracellular cholesterol accumulation (23), connecting LRP1 to another damaging factor of AKI – the intracellular accumulation of cholesterol. The local immune cells, in particular macrophages and T-cells, are involved in the pathophysiology of AKI by regulating renal repair, but may also induce the transition to CKD (24, 25). Macrophage LRP1 is a survival factor (26). In contrast, LRP1 agonism may increase the proportion of Treg cells (27), which is a beneficial effect in AKI recovery and the transition to CKD (25).

**Figure 1:**
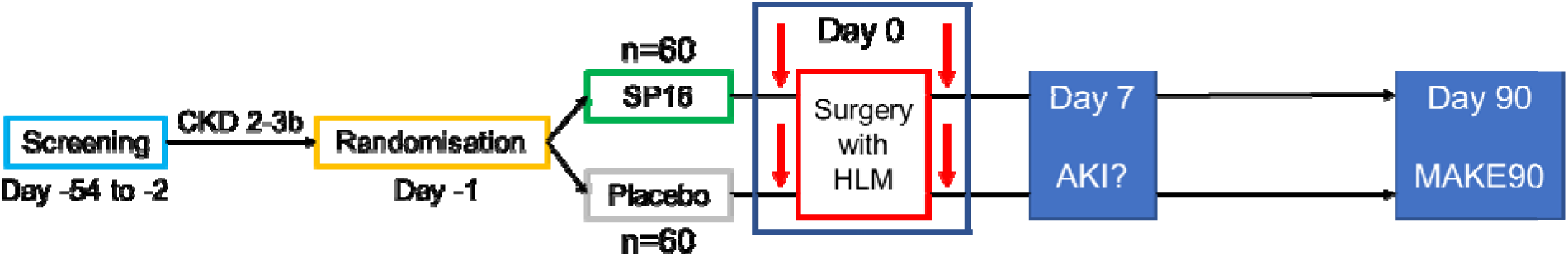
study flow diagram.

The LRP1 agonist SP16 was administered to healthy volunteers (phase I) (28) and to patients with myocardial infarction (phase I/II) (29) without showing any severe side effects. This clincial trial aims to evaulate the safety and the efficacy of SP16 to reduce the severity and rate of AKI as well as its transition to CKD/deterioration of existing CKD.

### Explanation for the choice of comparator {9b}

The double-blind, placebo-controlled design enables an unbiased evaluation of both efficacy and safety. The use of placebo is justified to allow assessment of the absolute efficacy and safety of SP16, given the absence of any approved therapeutic agents for reducing the risk of loss of kidney function in patients undergoing CS.

### Objectives {10}

The purpose of this trial is to investigate SP16 as potential renoprotective agent in CS patients.

The primary objectives are to evaluate the safety of SP16 administration and its ability to prevent CSA-AKI, as defined by KDIGO criteria, in participants with CKD 2-3b undergoing planned CS. Secondary objectives include evaluation of the effect of SP16 on the severity of CSA-AKI (according to KDIGO stages 1-3), the duration of CSA-AKI, the necessity and duration of RRT, the cardiac left and right ventricular functions, major adverse kidney events at day 90 (death, need for dialysis, sustained impaired renal function, MAKE90) and the incidence of AEs and SAEs.

## Methods: Patient and public involvement, and trial design

### Patient and public involvement {11}

The complexity and the requirements of the trial design excluded participation of lay persons.

### Trial design {12}

This investigator-initiated trial (IIT) is a phase IIa, randomised, double-blind, parallel group, single-centre clinical study that will evaluate the efficacy and safety of SP16 compared to placebo in 1:1 ratio in 120 participants with CKD G2-3b undergoing CS.

## Methods: Participants, interventions and outcomes

### Trial setting {13}

Single centre trial at the Universitätsklinikum Erlangen, Germany.

### Characteristics of the people who are needed for the trial

| Characteristic | The people we would expect to see included |
| --- | --- |
| Age | ≥18 y |
| Gender/Sex | Data from patients with coronary heart disease who are candidates for a coronary artery bypass graft (CABG) indicate a preponderance of the male gender (about 70:30), whereas for patients with diseased aortic valve or mitral valve no relevant gender difference is observed. Therefore, no measures to influence gender distribution in study enrollment will be taken. |
| Race, ethnicity and ancestry | n/a |
| Socioeconomic status | n/a |
| Geographic location | Germany |
| Other characteristics relevant to the trial | CKD Stage 2–3b with eGFR ≤90 and ≥30 ml/min/1.73 m<sup>2</sup> (according to 2021 CKD-EPI equation) known for ≥ 3 months prior to enrollment |

### Eligibility criteria for participants {14a}

Potential participants will be screened for eligibility at the CS department (out-patient and in-patient care) in accordance with the criteria outlined in Table 1, following the provision of written informed consent. The study will enroll adult patients scheduled for non-emergent coronary artery bypass graft (CABG) surgery, aortic valve surgery (aortic valve replacement or repair alone, with or without aortic root repair), mitral valve surgery or aortic/mitral valve surgery combined with CABG. To be eligible, patients must have already the diagnosis of CKD stage 2-3b. Key exclusion criteria include severe renal impairment, defined as an estimated glomerular filtration rate below 30 ml/min/1.73 m², emergent procedures, and current immunosuppressive treatment (see Table 2).

**Table 1:**
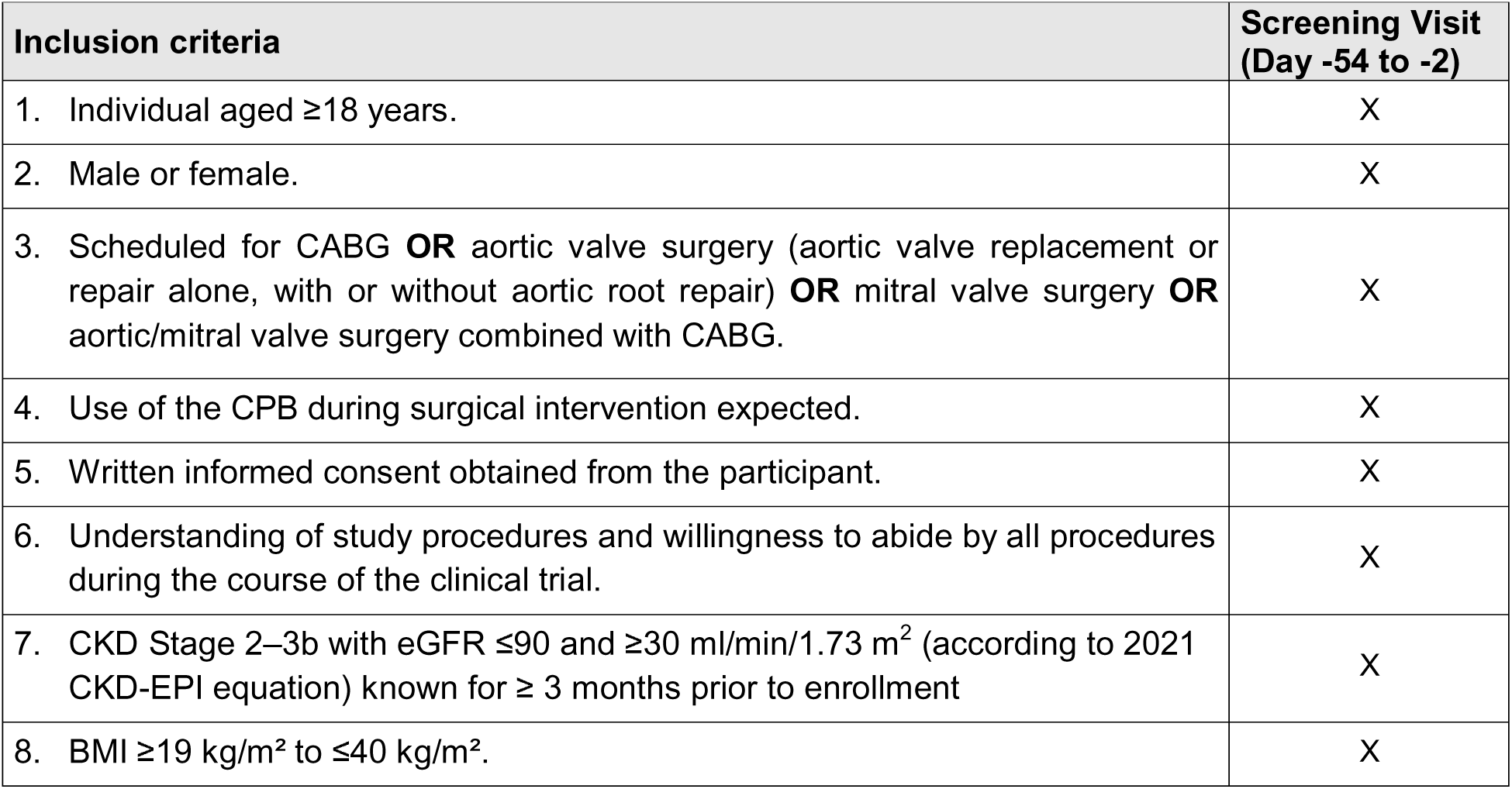
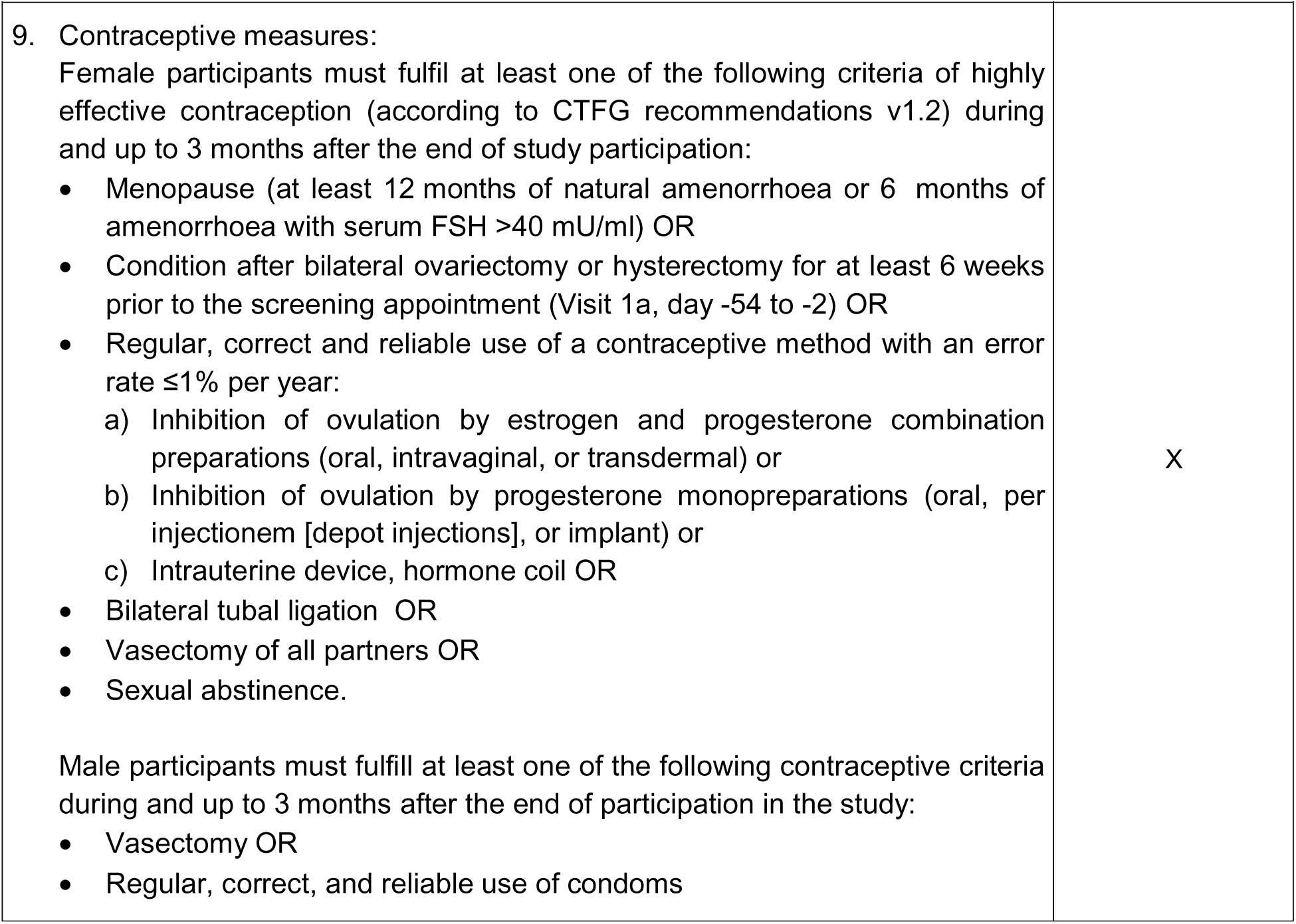
Inclusion criteria.

| Inclusion criteria | Screening Visit<br>(Day -54 to -2) |
| --- | --- |
| 1. Individual aged $\geq 18$ years. | X |
| 2. Male or female. | X |
| 3. Scheduled for CABG **OR** aortic valve surgery (aortic valve replacement or repair alone, with or without aortic root repair) **OR** mitral valve surgery **OR** aortic/mitral valve surgery combined with CABG. | X |
| 4. Use of the CPB during surgical intervention expected. | X |
| 5. Written informed consent obtained from the participant. | X |
| 6. Understanding of study procedures and willingness to abide by all procedures during the course of the clinical trial. | X |
| 7. CKD Stage 2–3b with eGFR $\leq 90$ and $\geq 30$ ml/min/1.73 m<sup>2</sup> (according to 2021 CKD-EPI equation) known for $\geq 3$ months prior to enrollment | X |
| 8. BMI $\geq 19$ kg/m<sup>2</sup> to $\leq 40$ kg/m<sup>2</sup>. | X |
| 9. Contraceptive measures:<br>Female participants must fulfil at least one of the following criteria of highly effective contraception (according to CTFG recommendations v1.2) during and up to 3 months after the end of study participation:<br>• Menopause (at least 12 months of natural amenorrhoea or 6 months of amenorrhoea with serum FSH >40 mU/ml) OR<br>• Condition after bilateral ovariectomy or hysterectomy for at least 6 weeks prior to the screening appointment (Visit 1a, day -54 to -2) OR<br>• Regular, correct and reliable use of a contraceptive method with an error rate $\leq 1\%$ per year:<br>a) Inhibition of ovulation by estrogen and progesterone combination preparations (oral, intravaginal, or transdermal) or<br>b) Inhibition of ovulation by progesterone monopreparations (oral, per injectionem [depot injections], or implant) or<br>c) Intrauterine device, hormone coil OR<br>• Bilateral tubal ligation OR<br>• Vasectomy of all partners OR<br>• Sexual abstinence.<br>Male participants must fulfill at least one of the following contraceptive criteria during and up to 3 months after the end of participation in the study:<br>• Vasectomy OR<br>• Regular, correct, and reliable use of condoms | X |

**Table 2:**
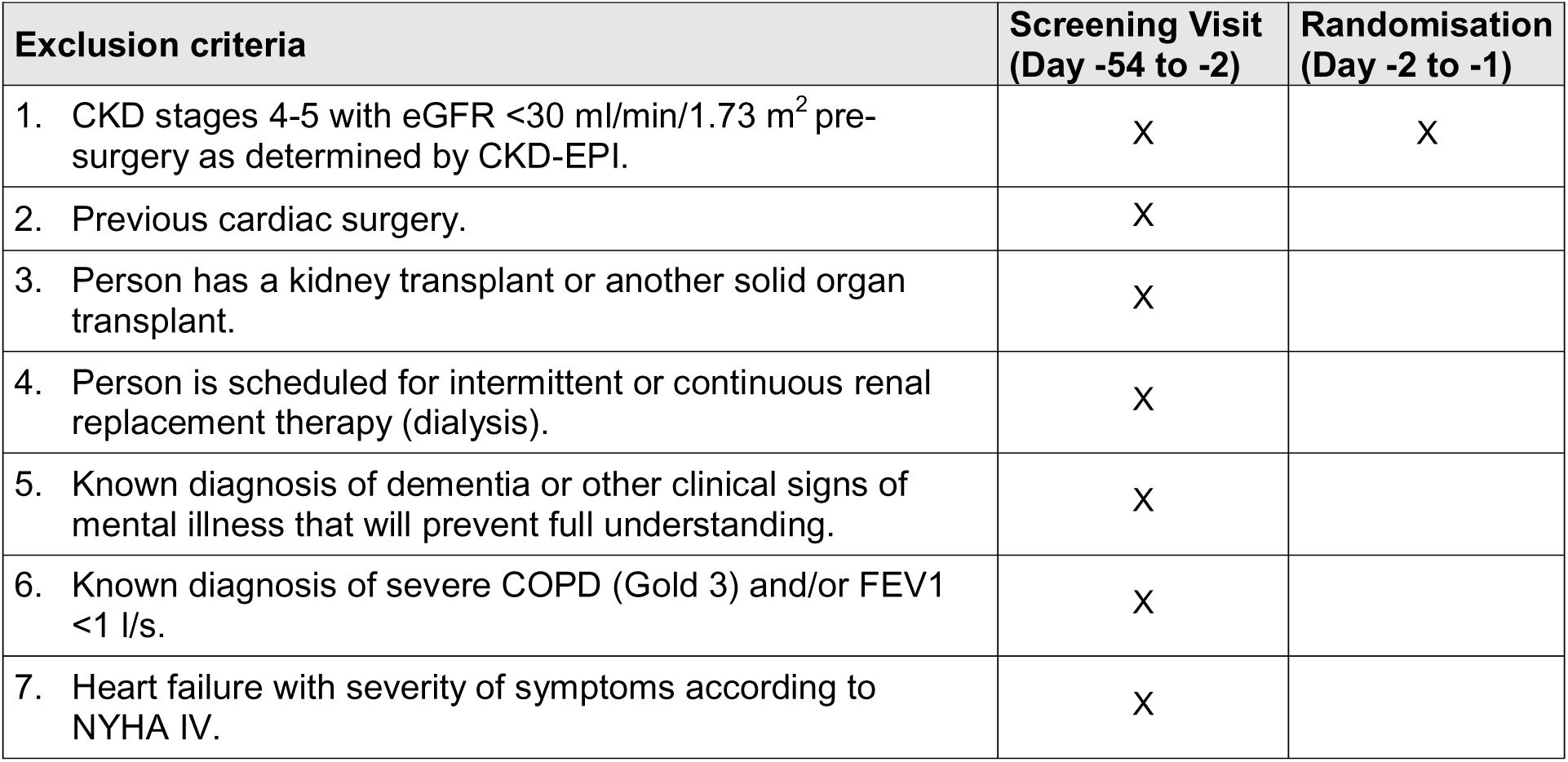
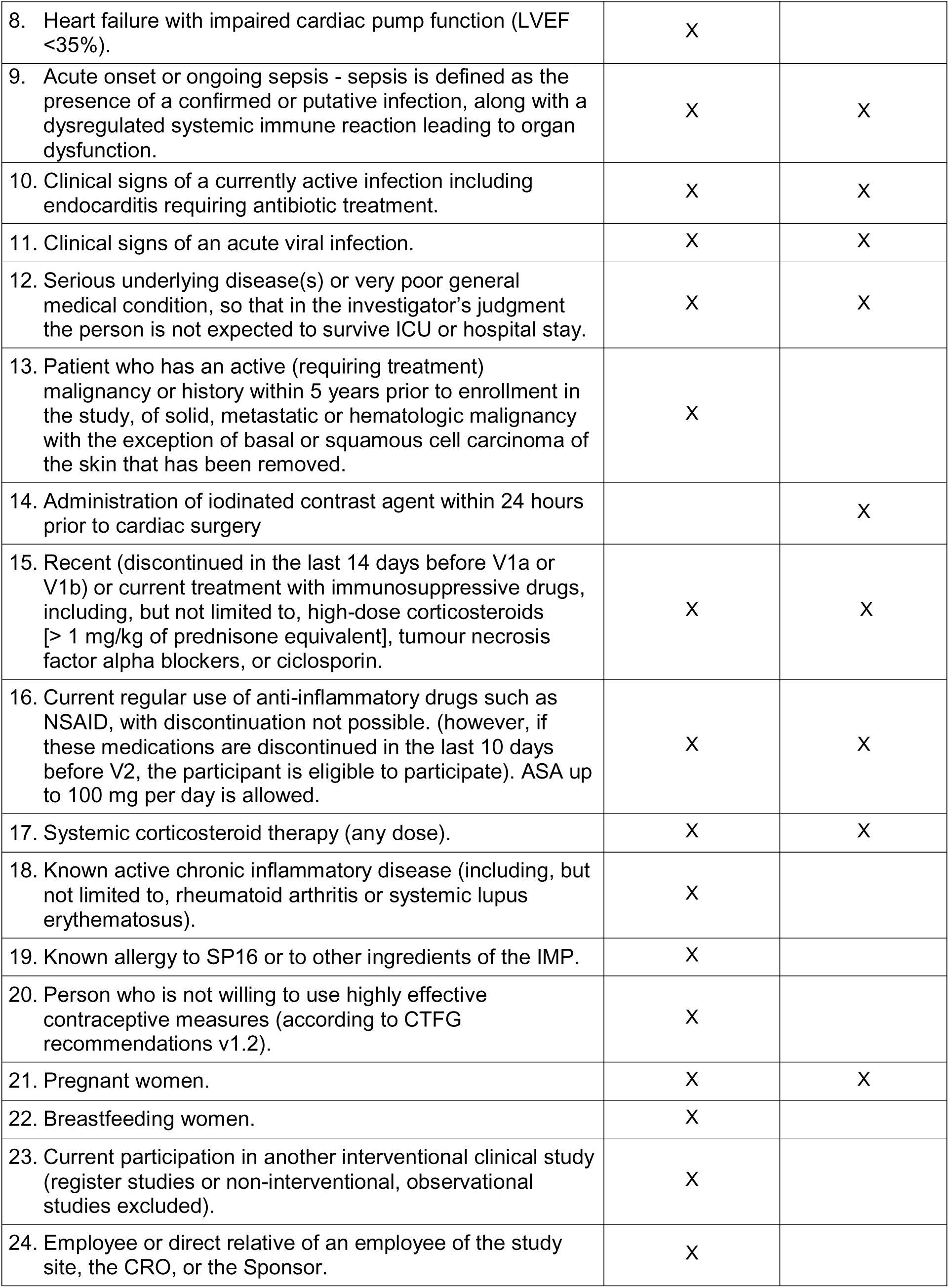
Exclusion criteria.

| Exclusion criteria | Screening Visit<br>(Day -54 to -2) | Randomisation<br>(Day -2 to -1) |
| --- | --- | --- |
| 1. CKD stages 4-5 with eGFR <30 ml/min/1.73 m<sup>2</sup> pre-surgery as determined by CKD-EPI. | X | X |
| 2. Previous cardiac surgery. | X |  |
| 3. Person has a kidney transplant or another solid organ transplant. | X |  |
| 4. Person is scheduled for intermittent or continuous renal replacement therapy (dialysis). | X |  |
| 5. Known diagnosis of dementia or other clinical signs of mental illness that will prevent full understanding. | X |  |
| 6. Known diagnosis of severe COPD (Gold 3) and/or FEV1 <1 l/s. | X |  |
| 7. Heart failure with severity of symptoms according to NYHA IV. | X |  |
| 8. Heart failure with impaired cardiac pump function (LVEF <35%). | X |  |
| 9. Acute onset or ongoing sepsis - sepsis is defined as the presence of a confirmed or putative infection, along with a dysregulated systemic immune reaction leading to organ dysfunction. | X | X |
| 10. Clinical signs of a currently active infection including endocarditis requiring antibiotic treatment. | X | X |
| 11. Clinical signs of an acute viral infection. | X | X |
| 12. Serious underlying disease(s) or very poor general medical condition, so that in the investigator's judgment the person is not expected to survive ICU or hospital stay. | X | X |
| 13. Patient who has an active (requiring treatment) malignancy or history within 5 years prior to enrollment in the study, of solid, metastatic or hematologic malignancy with the exception of basal or squamous cell carcinoma of the skin that has been removed. | X |  |
| 14. Administration of iodinated contrast agent within 24 hours prior to cardiac surgery |  | X |
| 15. Recent (discontinued in the last 14 days before V1a or V1b) or current treatment with immunosuppressive drugs, including, but not limited to, high-dose corticosteroids [> 1 mg/kg of prednisone equivalent], tumour necrosis factor alpha blockers, or ciclosporin. | X | X |
| 16. Current regular use of anti-inflammatory drugs such as NSAID, with discontinuation not possible. (however, if these medications are discontinued in the last 10 days before V2, the participant is eligible to participate). ASA up to 100 mg per day is allowed. | X | X |
| 17. Systemic corticosteroid therapy (any dose). | X | X |
| 18. Known active chronic inflammatory disease (including, but not limited to, rheumatoid arthritis or systemic lupus erythematosus). | X |  |
| 19. Known allergy to SP16 or to other ingredients of the IMP. | X |  |
| 20. Person who is not willing to use highly effective contraceptive measures (according to CTFG recommendations v1.2). | X |  |
| 21. Pregnant women. | X | X |
| 22. Breastfeeding women. | X |  |
| 23. Current participation in another interventional clinical study (register studies or non-interventional, observational studies excluded). | X |  |
| 24. Employee or direct relative of an employee of the study site, the CRO, or the Sponsor. | X |  |

### Eligibility criteria for sites and those delivering interventions {14b}

This study will be conducted within an established cooperation between the Department of Nephrology and Hypertension and the Department of Cardiac Surgery at the Universitätsklinikum Erlangen as the patient collective (subjects with chronic kidney disease undergoing elective cardiac surgery) links the expertise of both departments. Since it is a Phase II study, the IMP will be administered by physicians only.

### Who will take informed consent? {32a}

The investigator will inform potential participants about the nature of the study, including its risks and benefits, and will address any questions related to the study. Participants will be informed that participation is voluntary and that they may withdraw at any time. Prior to enrollment, participants must provide written informed consent in accordance with ICH guidelines, applicable local regulations (including 21 CFR 50), relevant privacy and data protection requirements, and the guidelines of the Institutional Review Boards (IRB), Research Ethics Boards (REB), or Independent Ethics Committees (IEC) overseeing the study site. The participant’s medical record must document that written informed consent was obtained before study enrollment, including the date of consent. The individual authorised to obtain informed consent must also sign the informed consent form (ICF). During the course of the study, participants will be reconsented using the most current relevant version of the ICF(s) as applicable. A signed copy of the ICF(s) will be provided to each participant.

### Additional consent provisions for collection and use of participant data and biological specimens {32b}

With separate consent from the participants, blood samples may be used for additional research conducted by the sponsor as an academic institution. This research may aim to improve the understanding of kidney injury, changes in renal function, the compound specific immune modulation, as well as the development of related or novel treatments or research methodologies.

## Intervention and comparator

### Intervention and comparator description {15a}

The active pharmaceutical substance SP16-3M, which has not yet been authorised in the EU, is used as the investigational medicinal product (IMP).

In the SP16 group, the IMP is injected subcutaneously at two different time points. The first administration will take place pre-surgically, in the area of the operating theatre, when the participant already is under anesthesia. The second administration will be performed post-surgically, 9±1 h after the first administration.

At each injection timepoint, 6 mg SP16, divided in 2x 3 mg (i.e. 2 ml), is injected subcutaneously at two different injection sites.

In the placebo group, placebo (dextrose 5%) is injected subcutaneously at the same timepoints as the IMP, at a fixed volume of 2 ml at two different injection sites.

For safety reasons, the study participants will be closely monitored for a period of approximately 1-2 h after completion of the injection. The monitoring is ensured by continuous intraoperative monitoring after the first IMP administration and by monitoring in the ICU after the second IMP administration.

### Criteria for discontinuing or modifying allocated intervention/comparator {15b}

If it is determined at randomisation (V1b) or baseline (V2), that circumstances have arisen in the participant since the screening visit (V1a) that lead to a violation of the inclusion or exclusion criteria, the participant will be withdrawn from the clinical trial before the application of any study medication. Premature discontinuation is documented in the eCRF as an unscheduled visit.

Other reasons for an individual participant to discontinue the study participation may include

- Serious violation of the protocol with impact on the reliability of the data.
- Any situation, disease, or clinically relevant change in laboratory values that, in the opinion of the investigator, would jeopardise the participant’s health and would not justify further participation in the clinical trial.
- Second CS within the observation period of 7 days after the index surgery (prior to the assessment of the primary efficacy endpoint).

After injection of at least one IMP, the participant should be followed until the final visit. In the case of serious protocol violations relevant to the quality of the data the participant should be excluded from the per protocol analysis or, if necessary, at the discretion of the Sponsor, from all other data analyses.

In accordance with the Declaration of Helsinki participants can withdraw from the clinical trial at any time at their own request without any disadvantages for their further treatment. The reason for the participant’s withdrawal from the trial will be documented in the eCRF, if communicated. All participants who withdraw prematurely from the clinical trial at their own request should be asked to undergo a final examination in an unscheduled visit at the earliest possible date, the results of which are documented in the eCRF. If they have already received the first administration of study medication, their data recorded until the time of withdrawal will be included in the statistical analysis.

The clinical trial should be halted temporarily by the Sponsor for safety review as soon as three SAEs suspected to be causally related to the IMP (meaning a Serious Adverse Reaction, SAR) occur. The Sponsor will first unblind the affected trial participants. If it is confirmed that the participants have been treated with SP16, recruitment of further patients will be suspended until the suspected causal relationship of the SAEs with SP16 has been clarified. The Sponsor shall notify the Member State concerned about the temporary halt and, where applicable, the resumption of the clinical trial within 15 d of the interruption and the resumption, respectively, via the EU portal, CTIS. Resumption after temporary halt constitutes a substantial modification requiring approval by the competent authority.

### Strategies to improve adherence to intervention/comparator {15c}

The study medication is administered subcutaneously twice in total, at each occasion by authorised study personnel, and not handed out to the participant. This ensures compliance to the study intervention to the highest degree.

### Concomitant care permitted or prohibited during the trial {15d}

The medicinal products listed below are permitted as concomitant medication for study participants during participation in the study, provided they are not listed in prohibited list below. The list contains medications usually administered after CS, but is not exhaustive. Other medications may be administered at the discretion of the attending physicians depending on clinical necessity.

- Intra- and post-surgical treatment in CS with CPB according to hospital standard. The therapy can be initiated prior to operation, in the intraoperative post-bypass period and continued during admission to the ICU or during the participant’s subsequent inpatient stay:

`o` For cardiovascular management to treat left and/or right ventricular dysfunction and/or vasoplegia (low vascular resistance): Inotropic agents and vasopressor (e.g. ephedrine, norepinephrine, epinephrine, dopamine, dobutamine, vasopressin, isoproterenol), beta-blocking agents (e.g. esmolol, metoprolol), calcium channel blockers (e.g. nicardipine), or direct vasodilators (e.g. hydralazine, nitroglycerin).
`o` Diuretics (e.g. for volume control, against potassium overload).
`o` For heart rhythm control to treat arrhythmias: e.g. beta-blocking agents, amiodarone.
`o` For analgesia: opioids, e.g. fentanyl, sufentanil, hydromorphone, morphine, remifentanil, methadone.
`o` For sedation: e.g. propofol, ketamine, dexmedetomidine, midazolam, lorazepam, diazepam, clonidin, lormetazepam.
`o` Oxygen.
`o` For hemostasis, coagulopathy: e.g. clotting factors, transfusion of red blood cells (RBC), platelets, or fresh frozen plasma (FFP), fibrinogen, cryoprecipitate, tranexamic acid (TXA), or desmopressin.
`o` For VTE prophylaxis: DOAC, low molecular weight heparin, unfractionated heparin (in advanced age patients [<80 y] who are at risk of renal insufficiency), argatroban.
`o` For systemic embolic events (SEE) prophylaxis if post-surgical CS-associated atrial fibrillation (AF) persists for more than 48 h: DOAC (e.g. apixaban), phenprocoumon (Marcumar^®^), warfarin, heparin, argatroban.
- Any prescribed medication for the treatment of chronic underlying diseases such as antihypertensive drugs, anti-arrhythmic drugs (e.g. beta-blockers, amiodarone), diuretics, antidiabetics, statins, anticoagulants, platelet inhibitors (e.g. ASA up to a dose of 100 mg/day, clopidogrel and its analoga).
- Any medication for the treatment of acute illnesses (e.g. antibiotics for intercurrent acute bacterial infections).
- Acetaminophen (paracetamol) or metamizole for headache, fever etc.
- Any non-prescription therapy for concomitant diseases.
- Necessary vaccinations as recommended by the STIKO.

The medicines can be used at the discretion of the attending physician.

The administration of the following drugs is prohibited during study participation:

- Corticosteroids at any dose during a period of 7 days after first dose of study medication.
- Interleukin inhibitors (IL-1 receptor antagonist anakinra, IL-6 antagonist tocilicumab).
- NSAIDs (e.g. ibuprofen, diclofenac, naproxen, ASA >100 mg/d).
- Aminoglycoside antibiotics.
- Further prohibited concomitant medication which is not listed above, but, at the discretion of the investigator, is necessary to ensure patient safety, might be included and will be documented accordingly.

If a clinical condition requires treatment with renally excreted drugs for which dose adjustment is recommended, or drugs known to alter renal function in a relevant manner, the dosage and timing of administration should be adjusted at the discretion of the investigator. Drugs should be documented accordingly.

### Ancillary and post-trial care {34}

After premature discontinuation of the clinical trial, the participant will receive standard treatment by the hospital’s attending physicians during the inpatient stay, followed by regular treatment by the attending general practitioner or specialist (e.g. cardiologist, nephrologist).

### Outcomes {16}

#### Primary endpoints

The efficacy endpoint is the change of the number of participants who develop CSA-AKI during hospital stay defined by KDIGO stage 1 or higher. The KDIGO guideline is the gold standard for assessment of AKI (30) and the defines the timeframe (AKI occurrence within 7 days after an event with potential AKI induction). The safety endpoint is the frequency of AEs and SAEs within 72 h after index surgery.

### Secondary endpoints

Secondary endpoints will evaluate the effect of SP16 on

- the severity of CSA-AKI (according to KDIGO stages 1-3).
- the duration of CSA-AKI.
- necessity of RRT.
- duration of RRT.
- the cardiac left and right ventricular function.
- the composite of major adverse kidney events (MAKE) that include death, need of dialysis, or sustained impaired renal function within 90±7 d after index surgery (MAKE90).
- the incidence of AEs and SAEs.

### Harms {17}

Injection site pain was reported by 15 of 16 patients in the SP16 arms and 3 of 6 patients in the placebo arm in the Phase I study (28). In both previous clinical studies (28, 29) no serious adverse reactions (SAR) have been observed following a single subcutaneous injection of SP16. From the clinical results to date it is concluded that SP16 is unlikely to have a significant effect on systemic safety in humans.

The clinical severity of an AE will be classified according to the Common Terminology Criteria for Adverse Events (CTCAE) version 5.0 as:

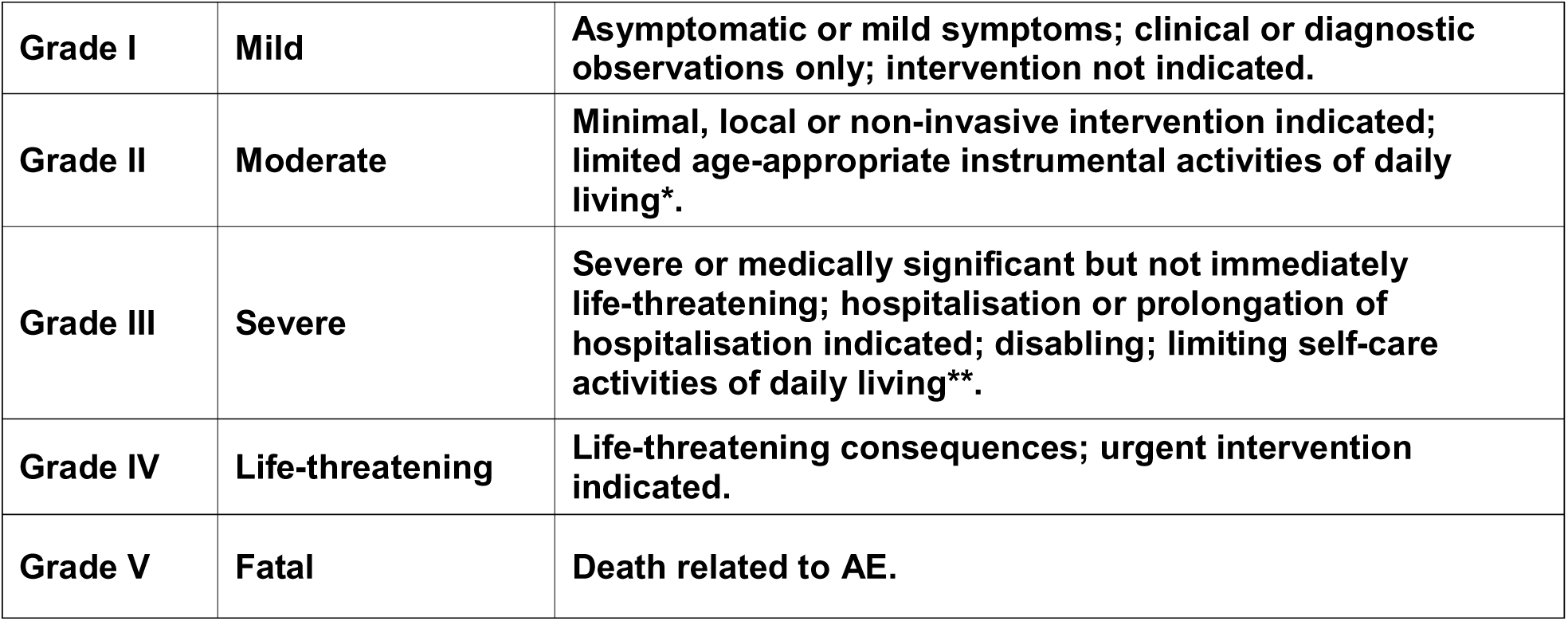

|  |  |  |
| --- | --- | --- |
| **Grade I** | **Mild** | **Asymptomatic or mild symptoms; clinical or diagnostic observations only; intervention not indicated.** |
| **Grade II** | **Moderate** | **Minimal, local or non-invasive intervention indicated; limited age-appropriate instrumental activities of daily living\*.** |
| **Grade III** | **Severe** | **Severe or medically significant but not immediately life-threatening; hospitalisation or prolongation of hospitalisation indicated; disabling; limiting self-care activities of daily living\*\*.** |
| **Grade IV** | **Life-threatening** | **Life-threatening consequences; urgent intervention indicated.** |
| **Grade V** | **Fatal** | **Death related to AE.** |

\*Instrumental activities of daily living (ADL) refer to preparing meals, shopping for groceries or clothes, using the telephone, managing money, etc.

*Instrumental activities 318 of daily living (ADL) refer to preparing meals, shopping for groceries or clothes, using the telephone, managing money, etc.

**Self-care ADL refer to bathing, dressing and undressing, feeding self, using the toilet, taking medications, and not bedridden.

### Participant timeline {18}

The study flow diagram (Figure 1) visualises the participant timeline.

The trial consists of 13 visits for each participant (PK samples only for 12 verum and 3 placebo patients to avoid unblinding):

· Visit 1a: Screening period of 52 days (day -54 to -2)

· Visit 1b (Randomisation, day -2/-1)

`o` The following tests only need to be performed if, in the opinion of the investigator, the values collected in V1a are too far apart to be used for randomisation of the patient.

`o` Pre-randomisation lab to support the review of selected exclusion criteria.

`o` Targeted physical examination.

`o` Recording of current concomitant medication.

`o` Randomisation.

· Visit 2 to 3: peri-operative treatment period (day 0)

`o` Visit 2 (Baseline, pre-surgery):

`o` Blood and urine samples for baseline laboratory tests including PK sample pre-surgery and before administration of the study medication (blank value) are taken directly before the 1^st^ administration of study medication (s.c. injection).

· Visit 3 (day 0 post-surgery)

`o` The 2 administration of study medication (s.c. injection) will be performed 9±1 h after the first administration.

`o` PK sample post-surgery (trough level) will be taken within 30 min before the 2^nd^ administration and further PK samples will be taken at timepoints 15±2 min, 30±3 min, 60±6 min, 90±9 min, and 180±18 min after 2^nd^ administration.

· Visit 4 to 11: post-treatment period during hospitalisation

`o` Visit 4 to 10: daily visits for 7 days post-surgery for the assessment of AKI according to the KDIGO guideline (primary endpoint at day 7)

`o` Visit 11: last study visit during hospitalisation (day 10±2 d or at discharge if earlier)

· Visit 12 and 13: Follow-up period until day 90 after surgery

`o` First follow-up visit at day 30±3

`o` Second follow-up and End of Study (EOS) visit at day 90±7

Study visits and assessments will be conducted in accordance with the schedules of assessments for the full study period (Table 3).

**Table 3:** Schedule of assessments.

|  | Screening | Randomisation | Baseline | OP | Hospitalisation |  |  |  |  |  |  |  |  |  |  |  |
| --- | --- | --- | --- | --- | --- | --- | --- | --- | --- | --- | --- | --- | --- | --- | --- | --- |
|  | Visit 1a | Visit 1b | Visit 2 |  | Visit 3 | Visit 4 | Visit 5 | Visit 6 | Visit 7 | Visit 8 | Visit 9 | Visit 10 | Visit 11 | Visit 12<br>(by phone) | Visit 13<br>/regular<br>EoS | unscheduled Visit<sup>1</sup><br>/Early<br>termination |
|  | Day -54 to -2 | Day -2 to<br>-1 | Day 0<br>pre-op |  | Day 0<br>post-op | Day 1 | Day 2 | Day 3 | Day 4 | Day 5 | Day 6 | Day 7 | Day 10<sup>2</sup>±2 | Day 30±3 | Day 90 ±7 |  |
| Informed Consent | ✓ |  |  |  |  |  |  |  |  |  |  |  |  |  |  |  |
| Inclusion/Exclusion criteria | ✓ | ✓ |  |  |  |  |  |  |  |  |  |  |  |  |  |  |
| Demographics | ✓ |  |  |  |  |  |  |  |  |  |  |  |  |  |  |  |
| Medical history | ✓ |  |  |  |  |  |  |  |  |  |  |  |  |  |  |  |
| Body-Mass-Index (BMI) | ✓ |  |  |  |  |  |  |  |  |  |  |  |  |  | ✓ | ✓ |
| Physical examination | ✓ |  |  |  |  |  |  |  |  |  |  |  | ✓ |  | ✓ | ✓ |
| Targeted physical examination |  | ✓<sup>3</sup> |  |  | ✓ | ✓ | ✓ | ✓ | ✓ | ✓ | ✓ | ✓ |  |  |  |  |
| Vital signs | ✓ | ✓ | ✓ |  | ✓ | ✓ | ✓ | ✓ | ✓ | ✓ | ✓ | ✓ | ✓ |  | ✓ | ✓ |
| ECG<sup>4</sup> | ✓ |  |  |  | ✓ | ✓ | ✓ | ✓ | ✓ | ✓ | ✓ | ✓ | ✓ |  | ✓ | ✓ |
| Echo | ✓ |  |  |  | ✓ | ✓ |  |  |  | ✓ |  |  |  |  | ✓ |  |
| Randomisation |  | ✓ |  |  |  |  |  |  |  |  |  |  |  |  |  |  |
| CPB parameters |  |  |  |  | ✓ |  |  |  |  |  |  |  |  |  |  |  |
| IMP Administration |  |  | ✓ |  | ✓ |  |  |  |  |  |  |  |  |  |  |  |
| Pre-Randomisation Lab |  | ✓<sup>3</sup> |  |  |  |  |  |  |  |  |  |  |  |  |  |  |
| Lab 1 - Safety Laboratory | ✓ |  | ✓ |  | ✓ | ✓ | ✓ | ✓ | ✓ | ✓ | ✓ | ✓ | ✓ |  | ✓ | ✓ |

<sup>1</sup> Unscheduled Visits may be performed more frequently if medically indicated at the investigators discretion or if a participant's participation is prematurely terminated
<sup>2</sup> or discharge from hospital whichever comes first
<sup>3</sup> tests only need to be performed if, in the opinion of the investigator, the values collected in V1a are too far apart to be used for randomisation of the patient.
<sup>4</sup> on ICU at least one ECG-recording should be taken after surgery (Day 0) and the next day (Day 1).

|  | Screening | Randomisation | Baseline | OP | Hospitalisation |  |  |  |  |  |  |  |  |  |  |  |
| --- | --- | --- | --- | --- | --- | --- | --- | --- | --- | --- | --- | --- | --- | --- | --- | --- |
|  | Visit 1a | Visit 1b | Visit 2 |  | Visit 3 | Visit 4 | Visit 5 | Visit 6 | Visit 7 | Visit 8 | Visit 9 | Visit 10 | Visit 11 | Visit 12<br>(by phone) | Visit 13<br>/regular<br>EoS | unscheduled Visit<sup>1</sup><br>/Early<br>termination |
|  | Day -54 to -2 | Day -2 to<br>-1 | Day 0<br>pre-op |  | Day 0<br>post-op | Day 1 | Day 2 | Day 3 | Day 4 | Day 5 | Day 6 | Day 7 | Day 10<sup>2</sup>±2 | Day 30±3 | Day 90 ±7 |  |
| Lab 2 -<br>Biomarkers<br>kidney function |  |  | ✓ |  | ✓ | ✓ |  | ✓ |  |  |  | ✓ |  |  |  |  |
| Lab 3 –<br>Biomarker<br>cardiac function |  |  | ✓ |  |  | ✓ |  |  |  |  |  | ✓ |  |  | ✓ | ✓ |
| Lab 4-accompanying<br>research |  |  | ✓ |  | ✓ | ✓ |  | ✓ |  |  |  | ✓ |  |  | ✓ |  |
| PK sampling |  |  | ✓ |  | ✓ |  |  |  |  |  |  |  |  |  |  |  |
| Urine output /<br>volume balance |  |  |  |  | ✓ | ✓ | ✓ | ✓ | ✓ | ✓ | ✓ | ✓ |  |  |  |  |
| Pregnancy test<br>if applicable in<br>WOCBP<sup>5</sup> | ✓ | ✓<sup>6</sup> |  |  |  |  |  |  |  |  |  |  |  | ✓ | ✓ | ✓ |
| AKI assessment<br>(KDIGO criteria) |  |  | ✓ |  | ✓ | ✓ | ✓ | ✓ | ✓ | ✓ | ✓ | ✓ | ✓ |  | ✓ | ✓ |
| Post-surgical<br>data |  |  |  |  |  |  |  |  |  |  |  |  | ✓ |  |  |  |
| Concomitant<br>medication | ✓ | ✓<sup>3</sup> |  |  | ✓ | ✓ | ✓ | ✓ | ✓ | ✓ | ✓ | ✓ | ✓ |  | ✓ | ✓ |
| Nephrotoxic<br>agents |  |  |  |  | ✓ | ✓ | ✓ | ✓ | ✓ | ✓ | ✓ | ✓ |  |  |  |  |
| AE assessment |  |  | ✓ |  | ✓ | ✓ | ✓ | ✓ | ✓ | ✓ | ✓ | ✓ | ✓ | ✓ | ✓ | ✓ |
| Study Diary<br>dispense/control |  |  |  |  |  |  |  |  |  |  |  |  | ✓ |  | ✓ | ✓ |

<sup>5</sup> according to CTFG [clinical trial facilitation group of the Heads of Medicines Agencies] recommendations for contraception and pregnancy testing in clinical trials v1.2 for WOCBP
<sup>6</sup> only if the test at V1a was carried out more than 72 h before the start of V2.

### Indicated laboratory work bundles include following laboratory tests

- Lab 1 (safety)

`o` Hematology (hemoglobin, hematocrit, red blood cells, MCH, MCV, MCHC, white blood cells, platelets, differential blood count [neutrophils/eosinophils/basophils/lymphocytes])
`o` Clinical chemistry (sodium, potassium, calcium, chloride, creatinine, eGFR, urea, anorganic phosphate, IL-6, IL-10, AST [GOT], ALT [GPT], γ-GT, bilirubin, LDH, alkaline phosphatase, lipase, lactate, creatine kinase [CK], creatine kinase-MB [CK-MB], troponine I (high-sensitive), total protein, albumin, CRP, uric acid)
`o` Coagulation (Thromboplastin time as Quick value [%], Thromboplastin time as INR, pTT [s])
`o` Urine (if available; specific gravity, pH, erythrocytes, leukocytes, bilirubin, proteins [quantitative], creatinine, albumin, IgG, transferrin, alpha-1-microglobulin will be determined in an urine sample [as far as appropriate volume available] by the local lab. Total protein and albumin will be analysed in relation to creatinine level [in gram or mmol]).
- Lab 2 (taken pre-surgery and 240±30 min, 24±1 h, 72±2 h, 7 d±6 h post-surgery) covers measurement of Cystatin C (serum) and NGAL (urine) as additional biomarkers for AKI assessment.
- Lab 3 determines NT-proBNP, a diagnostic biomarker of the cardiac function. Serum levels correlate well with the severity of acute heart failure.
- Lab 4 contains additional blood and urine samples for accompanying research program.

### Sample size {19}

This is a blinded, 1:1 randomised superiority trial with two parallel arms.

The primary efficacy endpoint is the occurrence of CSA-AKI within 7 days after surgery. This is a binary variable. The rate of CSA-AKI in the control arm is expected to be 50% (see (7), Table 1, row CKD 3). The rate of CSA-AKI in the treatment arm is expected to be 25%. Alpha is set to 5% and beta to 20%. A formula for a sample size estimation for a two-sided test can be found in equation 1 in (31).

Three interim analyses are planned (four analyses in total) according to a Group Sequential Method described by Wang and Tsiatis (see section 8.3 in (32)). This increases the sample size by a factor of 2.4% (see table 8.2.2 of (32)).

Drop-outs are not expected up to day 7 as the patients are in an ICU after CS.

This yields a sample size of 120 participants (60 per arm) and interim analyses at 30, 60 and 90 evaluable participants.

### Recruitment {20}

Between 2019 and 2024 the Department of Cardiac Surgery at the Universitätsklinikum Erlangen treated approximately 240 patients who suffered from CKD stages 2-3b and underwent aortic valve, mitral valve, CABG surgery, or a combination of these procedures using a CPB.

It is therefore assumed that the planned number of 120 patients for this study will be reached within the planned 38-month study period from the patient pool of the Departtment of Cardiac Surgery.

## Assignment of interventions: randomisation

### Sequence generation: who will generate the sequence {21a}

The randomisation sequence is created according to the Maximal Procedure described in (Berger et al., 2003) with the randomizeR software (Uschner et al., 2018). The randomisation will be stratified by gender.

### Sequence generation: type of randomisation {21b}

The participants are assigned to one of the two treatment arms, either IMP or placebo by means of 1:1 randomisation.

### Allocation concealment mechanism {22}

The random allocation sequence will be conducted with sealed, opaque envelopes according to the stratification by gender. Information on the random allocation sequence is only accessible for the pharmaceutical trial investigators, who will prepare the IMP.

### Implementation {23}

The randomisation list will be kept strictly confidential until unblinding and will not be accessible to anyone involved in the clinical trial with the exception of the responsible employees of the hospital pharmacy and the biobank CeBe of the Universitätsklinikum Erlangen.

## Assignment of interventions: blinding

### Who will be blinded {24a}

Study participants, investigators, and study centre staff involved in recruitment, treatment, and data collection are blinded to the study intervention. The unblinded study pharmacist or a suitably qualified individual responsible for preparing the study intervention for each participant will be informed. Access to the pharmacokinetic data record is restricted to a suitably qualified individual not involved in recruitment, treatment, or data collection.

### How will be blinding be achieved {24b}

The clinical trial will be conducted in a double-blind design. The investigators and study staff with patient contact at the trial site (e.g. study nurse) as well as the trial participants will be blinded. Further, the biostatistician and the person doing the pharmacovigilance will be blinded, too. The blinding will be maintained at least until the last participant last visit (LPLV) has been performed (or until the participant’s last visit if the participant dropped out earlier), data have been monitored and cleaned, and the data base has been closed.

### Procedure for unblinding if needed {24c}

To unblind participants, the study team/pharmacovigilance must check the sex of the patient to be unblinded. One complete set of emergency envelopes is sent to the trial site and one set is deposited with the person responsible for pharmacovigilance.

Each envelope will be labeled with the patient number, as provided by the EDC system. The envelope will list “male” and “female” according to stratification and the treatment group for each stratum.

Unblinding of individual study participants before this time will only take place in the following cases:

- The participant experiences a medical emergency the treatment of which requires mandatory knowledge of the identity of the administered study medication. In this case, the participant’s emergency envelope will be opened by the investigator or an appropriately authorised member of the study team. This person notes the date and time of opening on the envelope and signs it. The result of the unblinding is documented in the eCRF together with the date and time of opening.
- An SAE occurs in a participant that is assessed by the investigator or Sponsor as possibly causally related to the IMP and in addition assessed by the Sponsor as unexpected according the information given in the Reference Safety Information section of the valid version of the Investigatoŕs Brochure (IB). Prior to a potential suspected unexpected serious adverse reaction (SUSAR) report (or prior to interruption of the entire clinical trial), the participant’s emergency envelope is opened by the person responsible for pharmacovigilance of the clinical trial on behalf of the Sponsor. This person notes the date and time of opening on the envelope and signs it. The result of the unblinding is documented on the corresponding SAE Assessment Sheet.

## Data collection and management

### Plans for assessment and collection of outcomes {25a}

All required data will be entered by trained study site personnel into the eCRF through a secure electronic data entry system established by the internal Medical Centre for Information and Communication Technology at the University Hospital Erlangen. Data entry access is restricted to authorised, trained study staff with individual, password-protected credentials. Study-specific eCRF completion guidelines provide detailed instructions for data entry.

### Plans to promote participant retention and complete follow-up {25b}

After surgery and hospital discharge, participants will be followed up on days 30 and 90 after the first dose of the study intervention. Trained study staff will maintain contact with participants and provide reminders for scheduled follow-up visits at the study site.

### Data management {26}

All data collection and processing during this clinical trial will be performed in accordance with Regulation (EU) No. 2016/679 (General Data Protection Regulation, GDPR) and all applicable national data protection requirements.

This single centre clinical trial will be conducted at the Universitätsklinikum Erlangen, which is the Sponsor of this clinical trial and the “*controller”* in accordance with Art. 4(7) of the Regulation (EU) No. 2016/679.

The legal basis for the processing of the personal data is the informed consent of the participant in accordance with Art. 6(1)a and Art. 9(2)a GDPR in conjunction with §40b(6) Arzneimittelgesetz (AMG). Prior to trial participation each participant must be informed in written and by the investigator about the purpose and extent of the collection and use of personal data, particularly medical records and their rights according to GDPR. Furthermore, the participants are asked to give consent to direct access to their medical records for trial-related monitoring, auditing and inspections. Eligible participants who do not consent to the processing of data will not be included in the clinical trial.

Clinical monitors appointed by the Sponsor will visit the trial sites regularly and check the participants’ ICF. Data will not be used for analysis until the monitor has verified that the participants has given unambiguous consent to participate and to have data collected, transferred, and analysed in the trial. In the event of withdrawal of consent, the necessity for storing data will be evaluated. While the GDPR strengthens personal data protection rights, encompassing the right to access, rectification, and withdrawal of data, it also specifies the situations when restriction on those rights may be imposed. The withdrawal of informed consent should not affect the results of activities already carried out, such as the storage and use of data obtained on the basis of informed consent before.

### Confidentiality {33}

Within the means available to an academic institution, the Sponsor as well as the participating cooperation partners (“joint controllers”) and their contracted service providers (“processors”) are using several technical and organisational measures (TOMs) to protect the data.

In order to ensure data confidentiality and to prevent data misuse, the following TOMs are taken:

- Pseudonymisation of participants’ data.
- Encryption during data transmission in the eCRF with SSL (Secure Socket Layers) connections.
- Access control by central user administration.
- `o` Implementing access restriction to premises and data processing facilities where personal data are processed or biological samples are stored and processed (door can only be opened with key; administrative instructions on locking obligations).
- `o` Restricting access to protected personal data, data processing systems, study documents, patient files (e.g. password protection, service ID, log-in to clinical workstation, firewall, antivirus software).
- `o` Ensuring that only authorised and trained persons can access the data corresponding to their access authorisation (e.g. by means of a rights and roles concept, user accounts according to trial-specific delegation log).
- Dissemination control are measures to ensure that personal data cannot be processed without authorisation during electronic transmission or during their transport or storage on data carriers, and that it is possible to check and determine the location to which a transfer of personal data is intended (encrypted connection during data transmission, audit trail in the eCRF).
- Modification control as any change or correction to the eCRF is only permitted by authorised personnel in accordance with the provisions of ICH guideline E6 and must be traceable and recognisable (audit trail in EDC system, correction with date and signature on paper-based documents).
- Loss control by taking regular backups for digital data, audit trail and orderly filing of documents. Employees involved in the study are instructed by those responsible on

the duty of confidentiality and trained in the handling of personal data and samples.

- Quality management (QM) system based on Standard Operating Procedures (SOPs) and manuals for conducting the clinical trial.
- Strict separation between information technology (IT) infrastructures that process personal data and structures that do not.
- Strict requirements for the maintenance/updating of the systems, including requirements for the prompt installation of security-relevant patches and updates.

Strict guidelines for mobile working, including guidelines for the use of external storage

## Statistical methods

### Statistical methods for primary and secondary outcomes {27a}

The null hypotheses for the other endpoint variables are the same as for the primary efficacy variable: The variables and the study arms are independent. Appropriate statistical tests are chosen to test these hypotheses in the complete case population.

For the primary safety endpoint “Frequency of AE and SAE within 72 hours after index surgery”, the number of these events are counted per arm and compared with Poisson test. The number of participants with at least one of these events is also counted. This is a binary variable.

### Who will be included in each analysis {27b}

All randomised participants will be included in each analysis.

### How missing data will be handled in the analysis {27c}

Three additional analyses sets are defined:

- Complete case analysis (participants with available data on the primary efficacy variable only).
- Intention-to-treat analysis with imputation (all randomised participants; missing values of the primary efficacy variable are imputed).
- Per protocol analysis (participants with available data on the primary efficacy variable and no major protocol deviations only).

### Methods for additional analyses (e.g. subgroup analyses) {27d}

Intercurrent events are addressed with the treatment policy strategy (i.e. the occurrence of intercurrent event is irrelevant for the analysis) and with the principal stratum strategy (i.e. subgroup analysis for the administered drugs and generalised linear models with various drugs as covariates).

### Interim analyses {28b}

Three interim analyses are planned according to a Group Sequential Method described by Wang and Tsiatis (see section 8.3 in (32)).

### Protocol and statistical analysis plan {5}

Details of the study protocol including statistical considerations are available at <u>euclinicaltrials.eu</u> (EUCT number: 2025-522491-89-00). The full protocol and individual participant data will not be shared.

## Oversight and monitoring

### Composition of the coordinating centre and trial steering committee {3d}

There is no specific steering committee for this phase IIa trial.

### Composition of the data monitoring committee, its role and reporting structure {28a}

The Data and Safety Monitoring Board consists of three experts who are not involved in the operative conduct of the clinical trial and who are independent of the sponsor and funder. The DSMB evaluates the data on the IMP’s safety and tolerability and advises the Sponsor regarding the continuation, suspension, or early termination of the clinical study.

### Frequency and plans for auditing trial conduct {29}

In order to verify that the rights, safety, and well-being of participants are protected, that the reported data are reliable and robust, and that the conduct of this clinical trial follows the protocol, the ICH guideline E6, and all applicable national and international laws and regulations, the Sponsor shall adequately monitor the conduct of this clinical trial.

Prior to initiation of trial and enrollment of participants, the Sponsor or delegated monitor will review the protocol, CRF, procedures for obtaining informed consent, record keeping, and reporting of AEs with the investigator(s) and members of the investigating team.

The investigator provides the monitor with access to all trial-related original data and documents relevant for the monitoring of the trial.

The extent and nature of the monitoring is determined by the Sponsor. In principle, ICFs, adherence to inclusion/exclusion criteria, and documentation of SAEs including the proper recording will be reviewed as part of the monitoring process. In addition, the monitor will review trial records and directly compare them with source documents (source data verification, SDV), review proper handling of the IMPs, discuss the conduct of the trial with the investigator(s) and members of the investigating team, and verify that the clinical trial site remains acceptable.

Any missing data or data discrepancies will be communicated to the trial site for clarification/resolution. The investigator agrees to cooperate with the monitor to ensure that any problems detected in the course of the monitoring visits are addressed and resolved and therefore ensures the consistency of the trial with ICH guideline E6 and all applicable national and international laws and regulations.

### Protocol amendments {31}

Any amendments to the protocol must receive IRB/REB/IEC approval before being implemented, except for modifications needed to address an immediate risk to study participants. Additionally, the protocol and any significant amendments require health authority approval before initiation, unless the changes are necessary to prevent an immediate hazard to participants.

### Dissemination policy {8}

Study results will be reported in a clinical study report in accordance with the ICH E3 guideline and will be disclosed alongside periodic safety reports and clinical study summary reports, as required by applicable regulations. Study information and summary tables will be posted on the EU Clinical Trials Register (https://euclinicaltrials.eu/). Additionally, the study findings will be published in peer-reviewed journals.

## Discussion

The phase IIa study EASE-AKI aims to assess the efficacy and the safety of the LRP1 agonist SP16 in preserving renal function in CS patients at high risk for acute kidney injury. Patients undergoing CS with CPB are at high risk of developing CSA-AKI, particularly with pre-existing CKD (7). Various efforts are still being made to reduce the high incidence of CSA-AKI. To date, none of the measures tested have led to a relevant breakthrough, so there still is a high clinical need for renal protection in this vulnerable patient cohort.

It has been shown that an inflammatory reaction is a key factor in the development of AKI (24, 25). A promising therapeutic approach could be to trigger immunomodulatory cascades via LRP1. Experimental data showed both renal and myocardial protective effects by treatment with the LRP1-agonist SP16. SP16 was well tolerated in the two previous clinical trials (28, 29), one study already showing a trend towards a reduction of inflammatory response (29). Based on the physiological mechanism of action, promising preclinical and preliminary clinical data, SP16 will be investigated to evaluate preventive effects on the incidence of CSA-AKI by reducing the inflammatory reaction after hypoxia and reperfusion caused by the use of the CPB pump during cardiac surgery. In addition, this study will provide further safety data in this larger placebo-controlled study collective. Reduction of post-surgical end-organ damage to both kidney and heart is expected to lead to a significant decrease in the rate and severity of AKI and a faster or more pronounced myocardial recovery.

As already stated before (33) the key secondary endpoints, including MAKE90 are of additional importance besides the primary endpoint of the frequency of AKI in the participants. This extended surveillance of the participants allows the assessment of renal function during a stable post-surgical phase, thereby reducing the impact of perioperative and early postoperative confounding factors. In addition, this approach extends the classical short-term endpoint of AKI incidence as the primary endpoint in prevention studies. These considerations are further supported by recent findings from both interventional and observational AKI studies, showing that many CS patients who do not meet formal AKI criteria nonetheless develop irreversible and clinically meaningful declines in eGFR, while conversely, some patients who fulfill AKI criteria do not experience sustained renal impairment (34, 35).

The assessment of cardiac function and recovery is an additional important secondary endpoint, since the improvement of cardiac function after CS reduces the risk of AKI (36) whereas postoperative cardiac dysfunction increases the risk of CKD development (37). These findings underline the relevance of a potential synergistic effect of SP16 on both renal and cardiac protection in the treated patient cohort.

There are several limitations of this trial: Only one dose of SP16 divided in two administrations is investigated, however this dose is the same as the highest dose administered in previous SP16 studies. In addition, it is a single-centre study which limits the number of potential participants. However, this excludes the differences in surgical and postoperative intensive care procedures between different study sites which tends to provide larger treatment effects (38, 39). The study is not powered for secondary endpoints like MAKE90, but the selection of the primary endpoint with renal damage in the acute phase follows the recommendation for the design of a phase II AKI prevention trial (40).

In summary, this single centre phase IIa study EASE-AKI will assess the efficacy and the safety of the LRP1 agonist SP16 as potential novel treatment approach for the prevention of CSA-AKI.

## Trial status

The current protocol version number is 1.1 dated 2026 February 2. The first participant to be screened is planned for 2026 April 1.

## Abbreviations

AE: Adverse Event
ACEi: Angiotensin Converting Enzyme Inhibitor
ADL: Activities of Daily Living
AF: Atrial Fibrillation
AMG: Arzneimittelgesetz / German Medicinal Products Act
AKI: Acute Kidney Injury
ALT (GPT): Alanine Aminotransferase (glutamate-pyruvate-transaminase)
AR: Adverse Reaction
ARB: Angiotensin II Receptor-1 Blocker
ASA: Acetylsalicylic Acid
AST (GOT): Aspartate aminotransferase (glutamate-oxaloacetate-transaminase)
BfArM: Bundesinstitut für Arzneimittel und Medizinprodukte/Ferderal Institute for Drugs and Medical Devices
BMI: Body Mass Index
BNP: Brain Natriuretic Peptide
CABG: Coronary Artery Bypass Graft
CeBE: Central Biobank Erlangen
CK: Creatine Kinase
CK-MB: Creatine Kinase MB
CKD: Chronic Kidney Disease
CKD-EPI: Chronic Kidney Disease Epidemiology Collaboration
COPD: Chronic Obstructive Pulmonary Disease
CPB: Cardiopulmonary Bypass (heart-lung-machine)
CRP: C-reactive Protein
CSA: Cardiac Surgery-Associated
CTIS: Clinical Trial Information System
D: Day
Dl: Deciliter
DOAC: Direct Oral Anticoagulants
DSMB: Data and Safety Monitoring Board
ECG: Electrocardiogram
eCRF: Electronic Case Report Form
EDC: Electronic Data Capture
EDTA: Ethylene Diamine Tetraacetic Acid
EF: Ejection Fraction
eGFR: Estimated Glomerular Fitration Rate
EoS: End of Study
EU: European Union
FEV1: Forced Expiratory Volume per Second
FFP: Fresh Frozen Plasma
GOLD: Global Initiative for Chronic Obstructive Lung Disease
H: Hour
HF: Heart Failure
ICF: Informed Consent Form
ICH: International Conference on Harmonisation
ICU: Intensive Care Unit
IgG: Immunoglobuline G
IL: Interleukin
IMP: Investigational Medicinal Product
INR: International Normalised Ratio
IPD: Important Protocol Deviation
i.v.: IntravenouGOLD Global Initiative for Chronic Obstructive Lung Disease
h: Hour
HF: Heart Failure
ICF: Informed Consent Form
ICH: International Conference on Harmonisation
ICU: Intensive Care Unit
IgG: Immunoglobuline G
IL: Interleukin
IMP: Investigational Medicinal Product
INR: International Normalised Ratio
IPD: Important Protocol Deviation
i.v.: Intravenous
KDIGO: Kidney Disease: Improving Global Outcomes [organisation] (Global non-profit foundation; Belgium)
Kg: Kilogram
L: Liter
LDH: Lactate Dehydrogenase
LRP: Low-density Lipoprotein Receptor-related Protein
MAKE: Major Adverse Kidney Event
MCH: Mean Corpuscular Hemoglobin
MCHC: Mean Corpuscular Hemoglobin Concentration
MCV: Mean Corpuscular Volume
Mg: Milligram
MHH: Medizinische Hochschule Hannover/Hannover Medical School
Min: Minutes
Ml: Milliliter
Mmol: Millimol
N/A: Not Applicable
NGAL: Neutrophil Gelatinase-Associated Lipocalin
NSAID: Nonsteroidal Antiinflammatory Drug
NT-proBNP: N-Terminal Pro-B-Type Natriuretic Peptide
NYHA: New York Heart Association
PBMC: Peripheral Blood Mononuclear Cell
PK: Pharmacokinetics
PTT: Partial Thromboplastin Time
RRT: Renal Replacement Therapy
OP: Surgery
QM: Quality Management
SAE: Serious Adverse Event
SAR: Serious Adverse Reaction
s.c.: Subcutaneous
SCr: Serum Creatinine
SEE: Systemic Embolic Event
SOP: Standard Operating Procedure
STIKO: Ständige Impfkommission
SUSAR: Suspected Unexpected Serious Adverse Reaction
TOM: Technical and Organisational Measures
TXA: Tranexamic Acid
USAR: Unexpected Serious Adverse Reaction
V: Visit
VTE: Venous Thrombembolism
WOCBP: Women of Childbearing potential

## Data Availability

Details of the study protocol including statistical considerations are available at euclinicaltrials.eu (EUCT number: 2025-522491-89-00). The full protocol and individual participant data will not be shared.

https://euclinicaltrials.eu/

## Acknowledgements

We thank Dr. Stephanie Maas, Lena Teresa Dietz, and Dr. Armin Ströbel from the Centre for Clinical Studies at the University Hospital Erlangen for administrative contribution in preparing the protocol submission.

## Declarations

### Authors’ contributions {3a}

TJS, FH, and MS designed the study.

TJS and KB wrote the first draft of the manuscript. All authors critically reviewed and approved the final manuscript.

### Sources of funding and other support {7a}

The trial is funded by Serpin Pharma, Inc., Manassas, Virginia, USA.

### Availability of data and materials {6}

The sponsor and its delegates will have access to the full study database.

### Ethics approval and consent to participate {30}

The Ethics Committee of the Hannover Medical School (MHH) has approved the protocol, ICF and related documents within the official approval procedure by the German Federal Institute for Drugs and Medical Devices (BfArM). Written informed consent to participate will be obtained from all participants.

### Consent for publication

Not applicable.

### Competing interests {7b}

DA is an employee of Serpin Pharma, CG is founder and CEO of Serpin Pharma. All other authors declare that they have no competing interests.

1 Unscheduled Visits may be performed more frequently if medically indicated at the investigators discretion or if a participant’s participation is prematurely terminated

2 or discharge from hospital whichever comes first

3 tests only need to be performed if, in the opinion of the investigator, the values collected in V1a are too far apart to be used for randomisation of the patient.

4 on ICU at least one ECG-recording should be taken after surgery (Day 0) and the next day (Day 1).

5 according to CTFG [clinical trial facilitation group of the Heads of Medicines Agencies] recommendations for contraception and pregnancy testing in clinical trials v1.2 for WOCBP

6 only if the test at V1a was carried out more than 72 h before the start of V2.

## Notes

### Clinical Trial

EUCT number 2025-522491-89-00

### Clinical Protocols

https://euclinicaltrials.eu/

